# Long-term Outcomes of Prophylactic Total Gastrectomy in Patients with Hereditary Diffuse Gastric Cancer

**DOI:** 10.64898/2025.12.11.25342067

**Authors:** Hui Jun Lim, Maria O’Donovan, Claire Lamb, Sophie Samra, Nyarai Chinyama, Paul C Fletcher, Hisham Ziauddeen, Ju Yi Tai, Patrick Murphy, Richard H Hardwick, Massimiliano Di Pietro, Rebecca C Fitzgerald, J Robert O’Neill

## Abstract

**Importance:** Hereditary diffuse gastric cancer (HDGC) is predominantly due to germline *CDH1* variants and confers a high lifetime risk of diffuse gastric cancer (DGC). Prophylactic total gastrectomy (PTG) is thought to prevent DGC but has potential long-term functional, nutritional and quality-of-life (QOL) sequelae.

**Objective:** We report the clinical, pathological and long-term weight and QOL outcomes from a prospective cohort of patients with HDGC who underwent PTG.

**Design and Setting:** This is a single-centre prospective cohort study (Familial Gastric Cancer Study) of *CDH1* pathogenic variant carriers managed at Addenbrooke’s Hospital (2005–2024).

**Participants:** Patients with *CDH1* pathogenic variants and completed □2 QOL questionnaires were included.

**Intervention:** Patients who underwent PTG or endoscopic surveillance were included.

**Main Outcomes and Measures:** The clinical, pathological, weight and QOL outcomes for patients undergoing PTG (n=60) are reported. The QOL of patients undergoing endoscopic surveillance only (n=25) and surveillance followed by PTG were also reported. Longitudinal changes in QOL were analysed using linear mixed models with Bonferroni-Holm correction.

**Results:** Median age at PTG was 31 years (IQR 25–41); 75% underwent vagal-sparing PTG. The major complication rate was <2% with no operative mortalities and a median hospital stay of 8 days. Foci of intramucosal signet ring cell (SRC) carcinoma (pT1a) were seen in 88%, pT0 in 10% with a median number of SRC foci of 8. The disease-specific and overall-survivals was 100% with median follow-up of 9.7 years. Weight loss was universal, reaching ∼15% at 3 months and extending to 20% by 24 months. Partial weight regain was seen in those with normal pre-operative BMI between 2–5 years. QOL worsened markedly at 1–3 months but largely returned to baseline by 12–24 months and remained stable at baseline levels beyond 10 years. Younger patients (<35) had better role functioning (p=0.038) and men reported greater persistent appetite loss (p=0.030) following PTG.

**Conclusions and Relevance:** PTG can be undertaken with a low major complication rate, and is a curative treatment for HDGC. Although substantial early functional and weight effects occur, most QOL domains recover to baseline by 12–24 months and are sustained long-term with regular follow-up within a specialist multidisciplinary service.

**Key Points:** *Question:* What are the clinicopathological and long-term impact on weight and health-related quality of life outcomes for patients with hereditary diffuse gastric cancer who undergo prophylactic total gastrectomy?

*Findings:* In this study which included 60 patients with HDGC who underwent PTG, there was a lower major complication rate at <2% and the majority of patients at 88.0% had pT1a on gastrectomy specimens. Partial weight regain was seen in those with normal pre-operative BMI between 2 to 5 years and QOL worsened markedly at 1 to 3 months but largely returned to baseline by 12 to 24 months and remained stable at baseline levels beyond 10 years.

*Meaning:* PTG can be undertaken with a low major complication rate and most QOL domains return to baseline within 24 months and sustained in the long-term within the context of a specialist multidisciplinary service.

## Background

Hereditary diffuse gastric cancer (HDGC) syndrome is predominantly attributed to germline *CDH1* mutations.^1, 2^ Individuals with pathogenic germline *CDH1* mutations have a 33.5 fold increased risk of developing diffuse gastric cancer by the age of 30 and women with HDGC carry a 6 fold higher risk of lobular breast cancer.^3^ Although advances in genetic testing have improved identification of *CDH1* mutation carriers, the mechanisms by which *CDH1* and other genes drive HDGC remain poorly understood, limiting preventative strategies. Given the insidious onset and poor prognosis of advanced DGC, prophylactic total gastrectomy (PTG) remains the only definitive risk-reducing option.^4–7^ Current guidelines recommend PTG for pathogenic *CDH1* carriers, with endoscopic surveillance allowing individualised timing.^7, 8^ However, PTG may result in long-term functional and nutritional sequelae affecting health-related quality of life (QOL).^9^

As many individuals with HDGC are asymptomatic and may never develop cancer, understanding surgical risks and long-term outcomes is essential to informed decision-making.^10, 11^ Most QOL data following total gastrectomy are derived from patients treated for invasive disease, with limited follow-up or confounding cancer therapies.^12, 13^ Long-term studies in sporadic gastric cancer demonstrate persistent QOL impairment up to five years post-operatively, underscoring the need for extended assessment.^14^

To date, only a small number of retrospective studies have evaluated outcomes following PTG in HDGC, mostly limited to two years of follow-up.^15–19^ These suggest PTG is safe, with low major complication rates and resolution of most symptoms by two years. Only 1 study has assessed QOL beyond five years, reporting no significant differences over time.^19^ This highlights the need for long-term evaluation.

We therefore examined clinicopathological and long-term QOL outcomes in patients with HDGC participating in the prospective Familial Gastric Cancer cohort study and undergoing PTG at a national referral centre.

## Materials and Methods

### Patient Cohort

All patients with HDGC managed as part of a national referral service at Cambridge University Hospitals from 1st January 2005 to 31^st^ January 2025 and who were prospectively enrolled in the longitudinal Familial Gastric Cancer study (FGCS) were included. Patients in the FGCS provided written informed consent to the use of clinicopathological data and weight outcomes as well as completion of serial questionnaires. Research ethical approval for the FGCS was provided by the Anglia and Oxford Medical Research Ethics Committee (reference number 97/5/32) and subsequent amendments approved by the Cambridgeshire Research Ethics Committee. All patients consented to the study received a baseline endoscopy with high resolution white light endoscopy and narrow band imaging with magnification followed by targeted biopsies of visible lesions, with particular focus on pale areas, and random gastric biopsies were taken according to the Cambridge protocol. For patients who opted to undergo endoscopic surveillance, 6-monthly or annual surveillance was performed depending on endoscopy findings. Patients diagnosed with invasive disease on baseline endoscopy were excluded.

There were several factors considered in the decision-making for PTG, including age, co-morbidities, family history, burden of signet ring cell (SRC) foci on endoscopy and patient choice. All patients were discussed at a multidisciplinary meeting before proceeding with PTG. All patients underwent baseline endoscopy and extensive pre-operative counselling and prehabilitation with a multi-disciplinary team (MDT). Selective use of pre-operative CT was applied based on patient factors. Following PTG, regular assessments of patients’ nutritional status, diet, weight and symptoms were performed in conjunction with dietician input and oral supplementation or rarely nasojejunal enteral feeding if required due to nutritional failure. Surgical feeding jejunostomies were not placed at PTG.

Patients who continued surveillance underwent 6-monthly, annual or 2-yearly surveillance depending on various factors, including age, endoscopic findings and patients’ preference, following discussion with their gastroenterologist and with input as needed from the MDT.

### Surgical Approach and Peri-operative Protocol

All patients underwent PTG by the open approach via either midline or rooftop incision with 1 of 2 specialised upper gastrointestinal surgeons. Lymphadenectomy was D1 as per international guidance with some D0 cases in the early cohort and D2 selectively based on patient factors. The posterior vagal trunk and coeliac innervation was preserved where possible, but the anterior vagal trunk was ligated at the hiatus. An end-to-side circular stapled oesophagojejunostomy and side to side semi-mechanical jejuno-jejunostomy was performed in all cases with intra-operative confirmation of complete gastric mucosal resection before anastomosis. Cholecystectomy was undertaken if gallstones were identified pre-operatively. Analgesia was via epidural or surgically placed rectus-sheath catheters with intravenous patient-controlled opiate analgesia. Patients followed a standardised enhanced recovery pathway with a planned level 2 ward stay for 1 to 2 days and then managed on a level 0 ward. Nasogastric tubes were not used and surgical drains placed very selectively.

### Surgical and Post-operative Outcomes Assessment

Prospectively collected clinical, pathological and post-operative outcomes were maintained using a bespoke online database. Peri-operative outcomes included operative approach, duration, lymphadenectomy, duration of high dependency and total inpatient stay and post-operative complications. Complications within 30 post-operative days were reported according to the Gastrectomy Complications Consensus Group classification and using the Clavien-Dindo system with major complications defined as ≥Clavien-Dindo IIIa.^20, 21^ Post-operative mortality was regarded as within in-patient stay or 90 days. The interval from date of surgery to date of disease recurrence was defined as the disease-free interval and date of surgery to death from any cause as the overall survival interval, censoring at last follow-up date.

### Processing and Histopathological Assessment of Gastrectomy Specimen

Gastrectomy specimens were received fresh the stomach was opened along the greater curve and examined for macroscopically visible abnormalities or lesions. The proximal and distal resection margins were removed and fixed in 10% formalin. The anterior and posterior walls of the stomach were separated and pinned onto corkboards. The anterior and posterior walls were fixed for 24 hours in PAXgene**^®^**tissue FIX and 10% formalin respectively then unpinned and placed in PAXgene**^®^** tissue stabilizer for the anterior wall. The anterior and posterior stomach walls were divided into cardia, fundus, body, transition zone, antrum and pylorus. These were blocked entirely into cassettes for paraffin embedding. Alternate blocks were used for diagnostic and research purposes. Sections from diagnostic blocks were stained with haematoxylin and eosin to assess for in-situ, pagetoid signet ring lesions and SRC carcinoma foci. All cases were reviewed by upper gastrointestinal specialist pathologists. Histopathological data included size, location, number of in-situ SRC lesions and SRC foci, proximal and distal resection margins, completeness of excision of gastric mucosa, total number of lymph nodes harvested, number of positive lymph nodes and tumour-node-metastasis (TNM) staging summarised according to TNM staging of 8th edition of American Joint Committee on Cancer.^22^ Histological types were classified according to 2019 World Health Organization classification of tumours of the digestive system.^23^

### Weight Outcomes Assessment

Pre-operative weight was prospectively collected when patients were scheduled for surgery and used as the baseline to derive post-operative changes in weight and body mass index (BMI; defined as weight (kg)/ height^2^ (m)). Standard post-operative follow-up intervals included 2 and 6 weeks, 3, 6, 9 and 12 months and then 6 monthly intervals until the third post-operative year and annually following this or at sooner intervals if needed.

### Health-related Quality of Life Assessment

We prospectively assessed impact of PTG on QOL using European Organisation for Research and Treatment of Cancer (EORTC) QLQ-C30 and QLQ-STO22 questionnaires at post-operative months 1, 3, 12 and annually thereafter.^24, 25^ Raw questionnaire responses were used to derive scores using the scoring manuals.^26, 27^ For patients continuing surveillance, prospective questionnaire responses were requested at baseline endoscopy and annually at subsequent endoscopies. For analysis of QOL changes, the first questionnaire response during surveillance was taken to be the baseline (dynamic baseline). The QOL assessment immediately prior to surgery was taken as the baseline for relative analysis of changes following PTG.

### Statistical Methods

Continuous variables were summarised using median (interquartile or total range as described), and categorical variables by frequency (%). The scores were summarised using mean (95% confidence intervals). To account for inter-subject variance in baseline and subsequent QOL measures, the percentage change from baseline was used for subsequent statistical analyses. Longitudinal QOL changes were analysed using linear mixed models with time, age grouped as <35 or >35, sex and vagal_sparing status as fixed effects and patient as a random effect.^28^ First_order autoregressive covariance was selected for residuals. Bonferroni-Holm correction accounted for multiple comparisons. A random intercept for center was included to account for within-center clustering. The model with the lowest BIC value was used to ensure robustness of the model. A two-sided P<0.05 indicated statistically significant. Analyses were performed in the R statistical environment, version 4.4.1 (R Foundation) and Jamovi, version 2.7.^29, 30^

## Results

### Patient Characteristics and Clinicopathological Outcomes

The patient characteristics, clinical and histopathological outcomes of patients who underwent endoscopic surveillance and PTG are shown in Table 1. An overview of the clinical pathway is shown in Figure 1. A total of 60 patients underwent PTG with median age at surgery of 31 years old (25, 41) and 45% (27/60) were female. Within the PTG cohort, there were 24 patients who initially opted for endoscopic surveillance and underwent at least 1 surveillance endoscopy and proceeded to PTG with a median number of 3 surveillance endoscopies over a median of 18 months. All patients having PTG had pathogenic *CDH1* variants. There were 3 patients who underwent endoscopic surveillance followed by PTG who completed at least 2 pre-operative and 2 post-operative QOL questionnaires.

**Figure 1:**
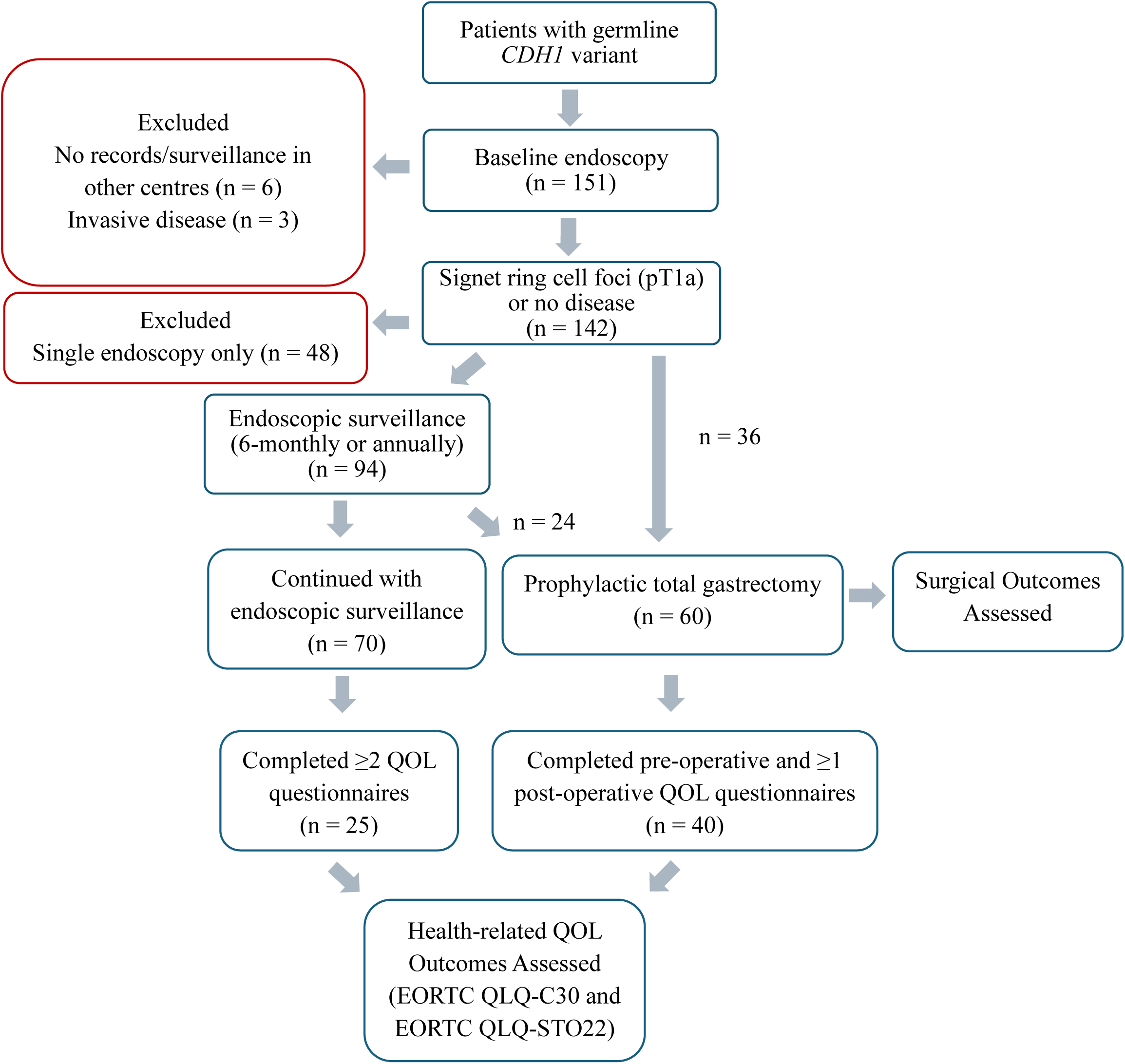
Flowchart showing Pathway of Patients with Hereditary Diffuse Gastric Cancer who underwent Endoscopic Surveillance and/or Prophylactic Total Gastrectomy.

**Table 1:**
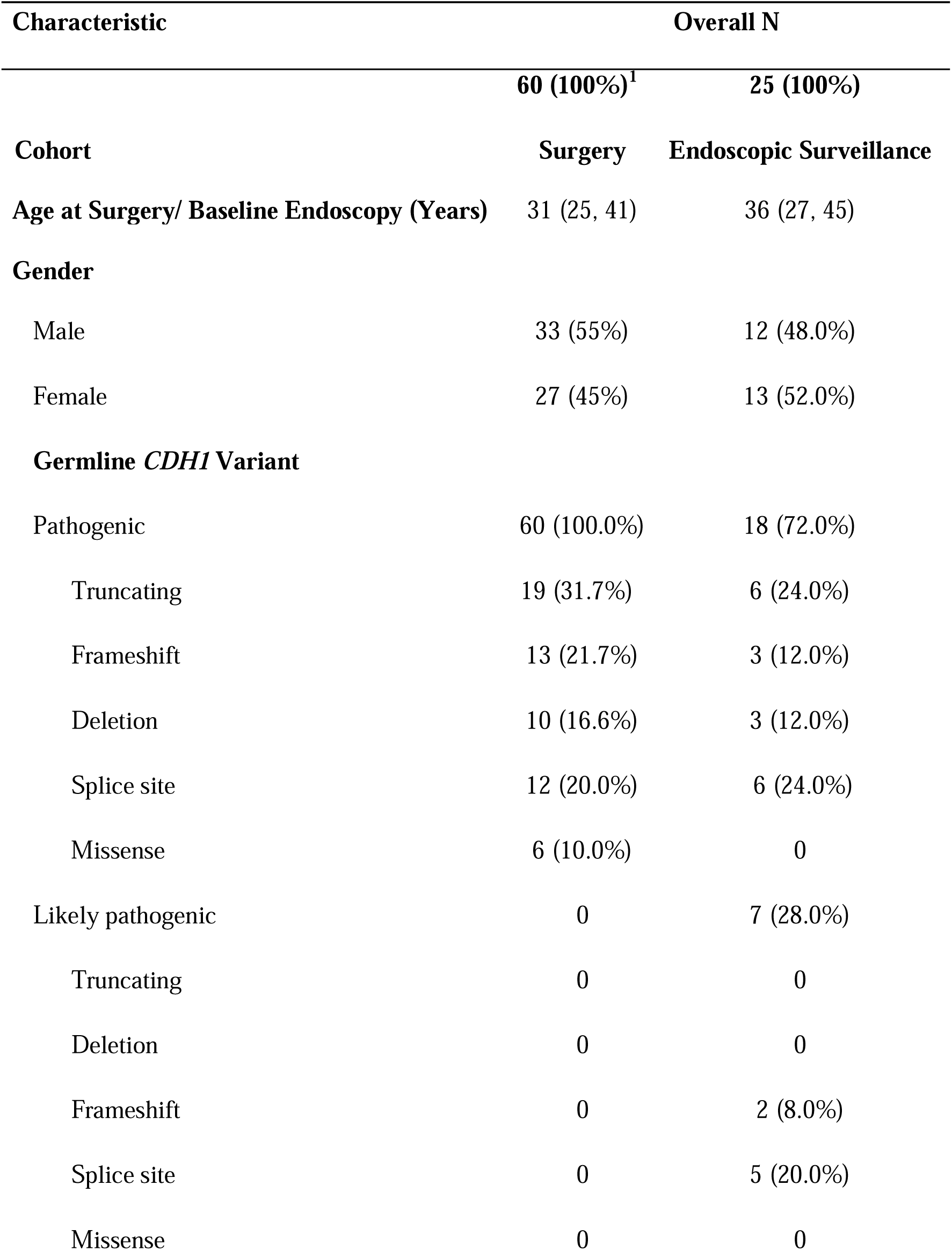
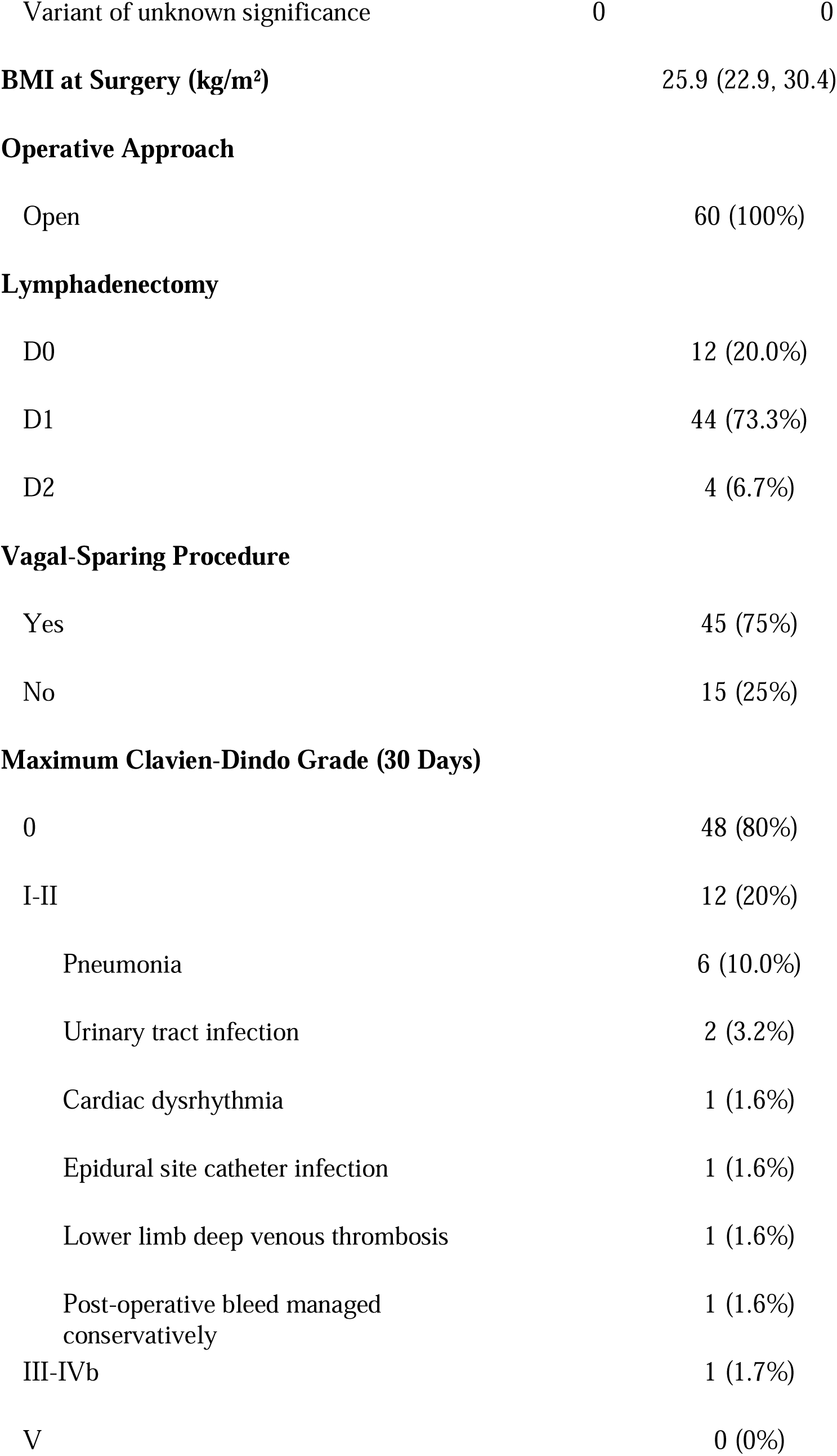
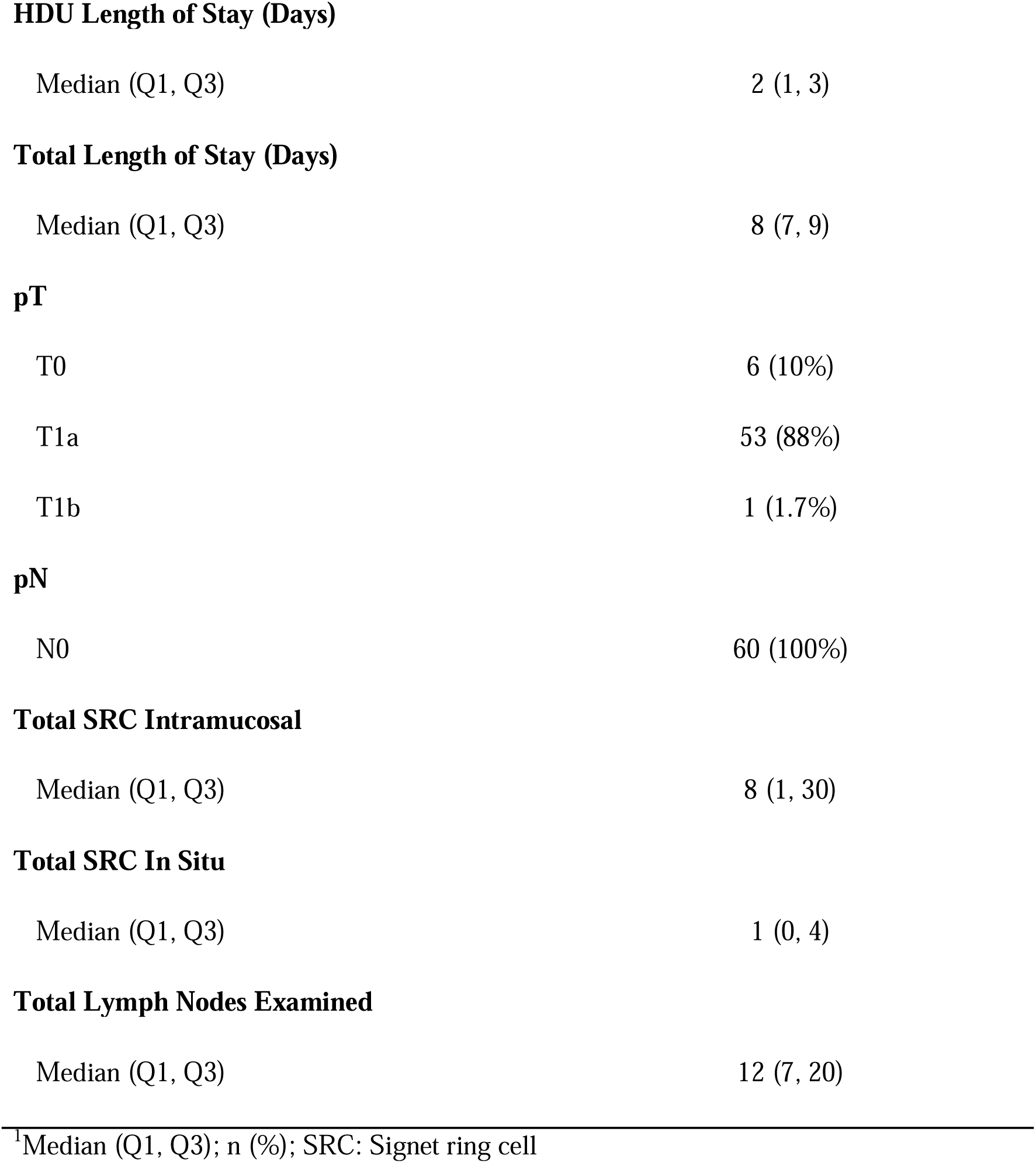
Summary of Patient Characteristics of Surveillance Cohort with QOL Data and.

| Characteristic | Overall N |  |
| --- | --- | --- |
|  | 60 (100%)<sup>1</sup> | 25 (100%) |
| **Cohort** | **Surgery** | **Endoscopic Surveillance** |
| **Age at Surgery/ Baseline Endoscopy (Years)** | 31 (25, 41) | 36 (27, 45) |
| **Gender** |  |  |
| Male | 33 (55%) | 12 (48.0%) |
| Female | 27 (45%) | 13 (52.0%) |
| **Germline *CDH1* Variant** |  |  |
| Pathogenic | 60 (100.0%) | 18 (72.0%) |
| Truncating | 19 (31.7%) | 6 (24.0%) |
| Frameshift | 13 (21.7%) | 3 (12.0%) |
| Deletion | 10 (16.6%) | 3 (12.0%) |
| Splice site | 12 (20.0%) | 6 (24.0%) |
| Missense | 6 (10.0%) | 0 |
| Likely pathogenic | 0 | 7 (28.0%) |
| Truncating | 0 | 0 |
| Deletion | 0 | 0 |
| Frameshift | 0 | 2 (8.0%) |
| Splice site | 0 | 5 (20.0%) |
| Missense | 0 | 0 |
| Variant of unknown significance | 0 | 0 |
| **BMI at Surgery (kg/m²)** | 25.9 (22.9, 30.4) |  |
| **Operative Approach** |  |  |
| Open | 60 (100%) |  |
| **Lymphadenectomy** |  |  |
| D0 | 12 (20.0%) |  |
| D1 | 44 (73.3%) |  |
| D2 | 4 (6.7%) |  |
| **Vagal-Sparing Procedure** |  |  |
| Yes | 45 (75%) |  |
| No | 15 (25%) |  |
| **Maximum Clavien-Dindo Grade (30 Days)** |  |  |
| 0 | 48 (80%) |  |
| I-II | 12 (20%) |  |
| Pneumonia | 6 (10.0%) |  |
| Urinary tract infection | 2 (3.2%) |  |
| Cardiac dysrhythmia | 1 (1.6%) |  |
| Epidural site catheter infection | 1 (1.6%) |  |
| Lower limb deep venous thrombosis | 1 (1.6%) |  |
| Post-operative bleed managed conservatively | 1 (1.6%) |  |
| III-IVb | 1 (1.7%) |  |
| V | 0 (0%) |  |

**HDU Length of Stay (Days)**
|  |  |
| --- | --- |
| Median (Q1, Q3) | 2 (1, 3) |

**Total Length of Stay (Days)**
|  |  |
| --- | --- |
| Median (Q1, Q3) | 8 (7, 9) |

**pT**
|  |  |
| --- | --- |
| T0 | 6 (10%) |
| T1a | 53 (88%) |
| T1b | 1 (1.7%) |

**pN**
|  |  |
| --- | --- |
| N0 | 60 (100%) |

**Total SRC Intramucosal**
|  |  |
| --- | --- |
| Median (Q1, Q3) | 8 (1, 30) |

**Total SRC In Situ**
|  |  |
| --- | --- |
| Median (Q1, Q3) | 1 (0, 4) |

**Total Lymph Nodes Examined**
|  |  |
| --- | --- |
| Median (Q1, Q3) | 12 (7, 20) |

<sup>1</sup>Median (Q1, Q3); n (%); SRC: Signet ring cell

The median operative time for PTG was 149 minutes (IQR 131, 234), and 45 patients (75.0%) underwent posterior vagal-sparing total gastrectomy. A total of 44 (73.3%) underwent D1 lymphadenectomy. A patient had a major complication with an anastomotic leak from the jejuno-jejunal anastomosis diagnosed on post-operative day 9 requiring re-operation (Clavien-Dindo IIIb). There were no post-operative mortalities. The median length of high dependency stay was 2 days (range 0 to 5) with a median inpatient stay of 8 days (range 5 to 17).

### Histopathological Outcomes

All patients had complete resection of gastric mucosa and R0 resection. The final stage was pT1bN0M0 in 1 patient (1.7%) which was due to superficial submucosal invasion in 1 lesion in the PTG specimen. This represents 0.05% (1/1697) of all histopathological lesions diagnosed on gastrectomy specimens. The final pathological stage was pT1aN0M0 in 53 patients (88%) and pT0N0M0 in 6 (10%) reflecting the high rate of SRC foci in patients with pathogenic *CDH1* mutations. The median number of SRC foci was 8 but there was a wide range from 0 to 273 reflecting a wide heterogeneity in this group and the choice of timing for PTG. The median number of lymph nodes harvested was 12 (range 0 to 54). No peri-neural or lympho-vascular invasion was reported. Disease-specific survival (DSS) and overall survival were 100% with median follow-up of 9.7 years (Supplementary Figure 1).

### Assessment of Long-term Weight Changes Following PTG

The post-operative weight and BMI of 57 patients was collected during follow-up. Post-operative weight loss was rapid and universal reaching 15% of baseline at 3 post-operative months. This reached a plateau of 15% to 20% total body weight loss between 12 and 24 post-operative months (Figure 2a). When stratified by pre-operative BMI, all patients lost weight by 12 post-operative months, but the greatest absolute loss occurred in patients with a higher pre-operative BMI (Figure 2b). Furthermore, in patients with a normal pre-operative BMI, weight gain was seen between 2 and 5 years post-operatively with most in a BMI range 20 to 25 kg/m^2^.

**Figure 2:**
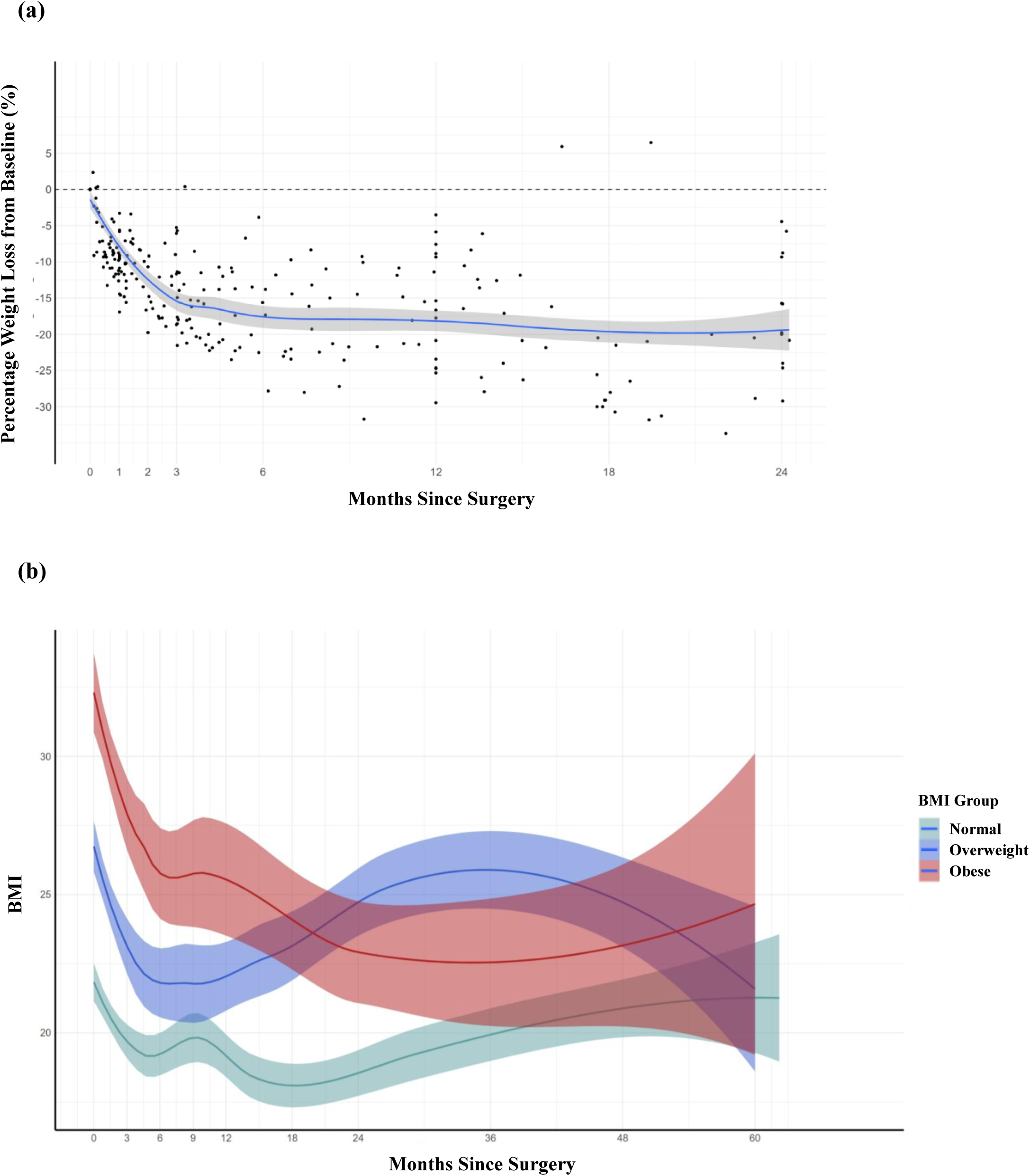
**a:** Weight Loss from Pre-operative Baseline (%) following Prophylactic Total Gastrectomy. **b:** Change in Body Mass Index (BMI) following Prophylactic Total Gastrectomy in Patients Grouped by Pre-operative BMI with Normal (BMI 20 to 25), Overweight (BMI 25 to 30) and Obese (BMI >30) Groups. Lines represent best-fit lines and error bars are 95% confidence intervals of the best-fit line.

### Health-related Quality of Life Outcomes

A summary of the scores in various functional domains and symptom scales in the surveillance cohort is shown in Figure 3. As anticipated, beyond small variances in symptom and functional domain scores, all outcomes were stable over 3 years.

**Figure 3:**
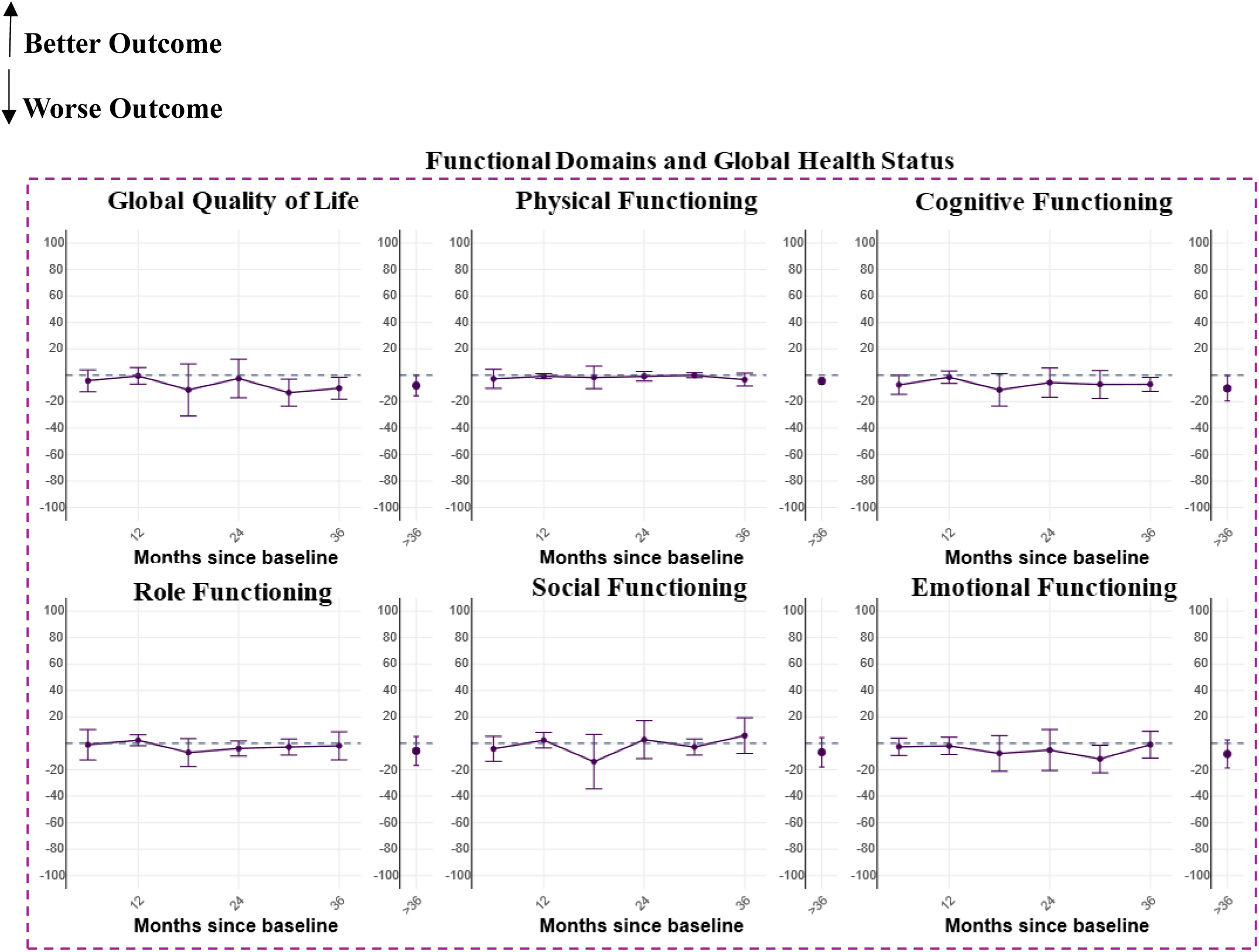
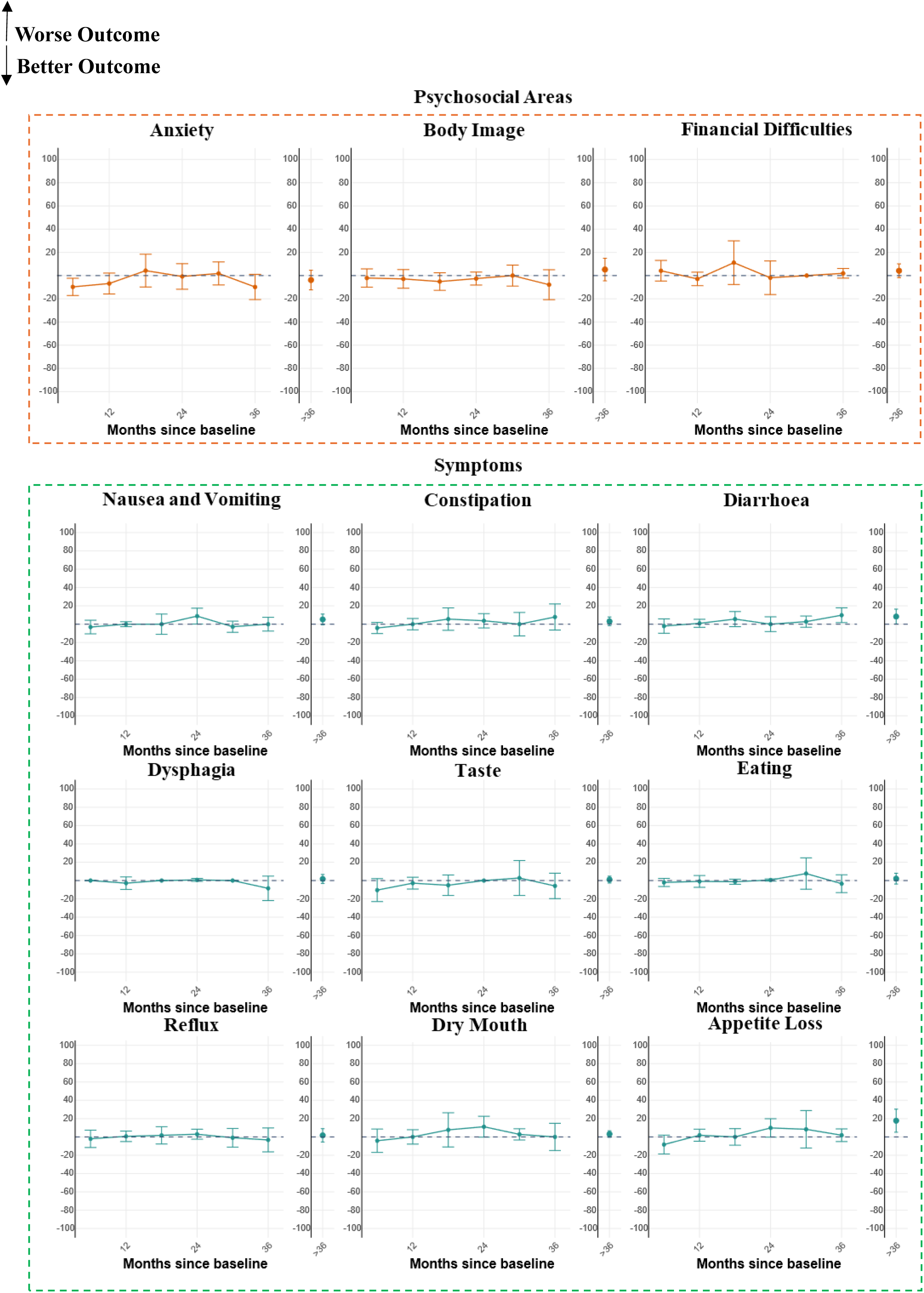
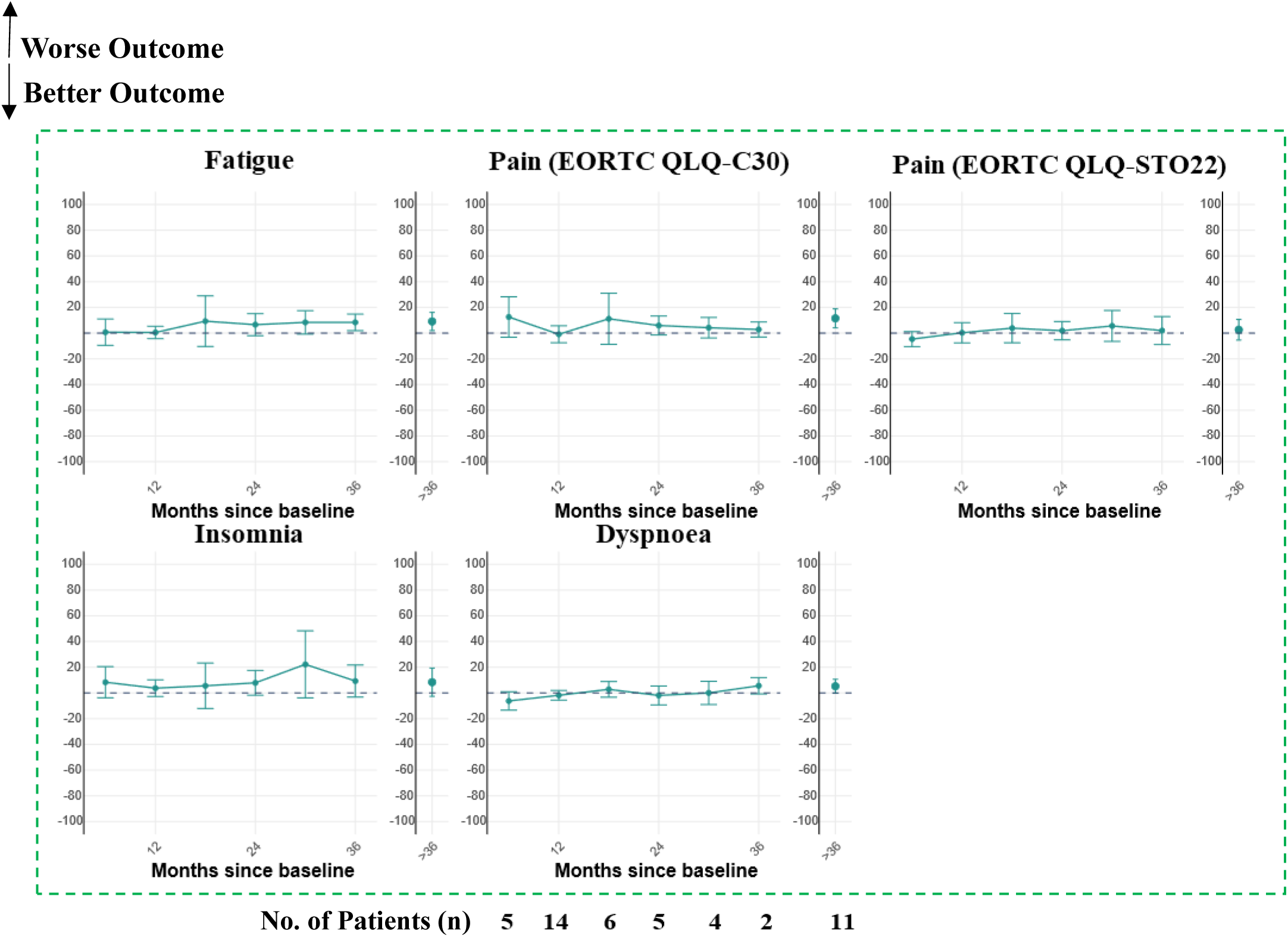
Change in EORTC QLQ-C30 and EORTC QLQ-STO22 Scores (mean ± 95% confidence interval) from Dynamic Baseline over Time (months) in Endoscopic Surveillance Cohort. The number of patients included at the various timepoints during endoscopic surveillance is shown below the bottom row of graphs and is representative for all the graphs.

**Figure 4:**
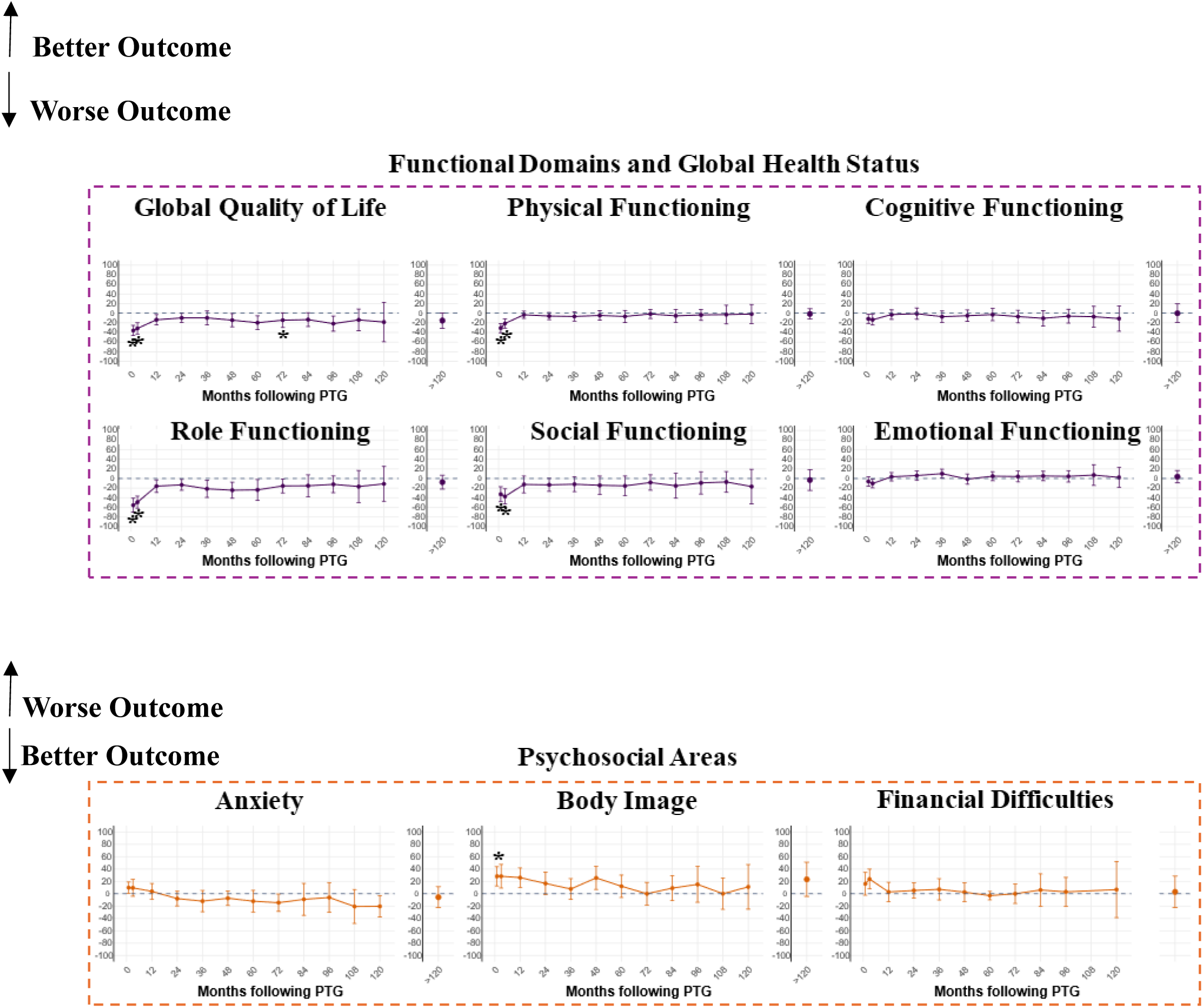
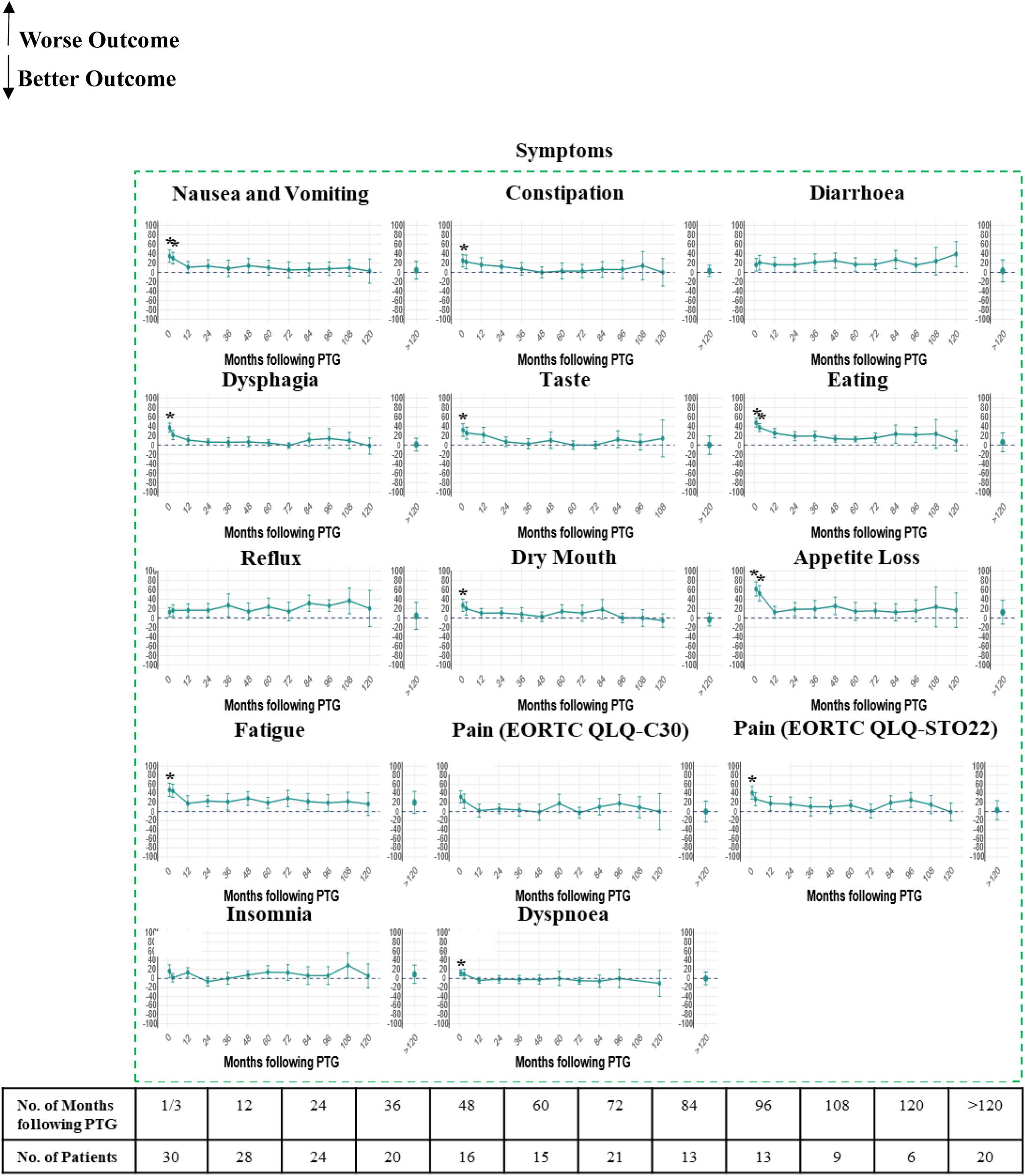
Change in EORTC QLQ-C30 and EORTC QLQ-STO22 Scores (mean ± 95% confidence interval) from Baseline (pre-surgery) over Time in Patients following Prophylactic Total Gastrectomy (* = significant change from baseline (P<0.05)). The number of patients included at the various timepoints following PTG is shown below the bottom row of graphs and is representative for all the graphs.

Changes in QOL from pre-operative baseline until after the first post-operative decade are shown in Figure 5. Significant changes were seen with a high rate of most symptoms and lower functional status across most domains at 1 and 3 post-operative months. Most changes had resolved to baseline levels by 12 or up to 24 months. Strikingly, in long-term follow-up from 2 to beyond 10 years, patients did not report significant different functional levels from baseline. Similarly, in symptom scores, recovery was close to baseline at 12 or up to 24 months. A trend was noted in more prevalent symptoms relating to appetite loss, eating difficulties, diarrhoea, reflux and fatigue although no significant difference from baseline could be observed at 1 years onwards to >10 years (Figure 3). A trend was also noted in reduced anxiety over time although this did not reach significance compared to baseline levels.

The changes in QOL outcomes for patients undergoing PTG were compared according to age at time of surgery (Supplementary Figure 2), gender (Supplementary Figure 3) and posterior vagal-sparing or non vagal-sparing surgery (Supplementary Figure 4). There were no significant differences in majority of functioning domains and symptoms within these groups. However, younger patients (<35) had significantly better role functioning **(p-value = 0.038)**. Most symptom changes were similar between genders however, men experienced significantly more appetite loss compared to women and this persisted in the long-term (**p-value = 0.030**). A summary of key findings is shown in Supplementary Figure 5.

We next sought to examine absolute QOL score changes in the same patients directly from surveillance through PTG and then into follow-up where 3 patients were included (Supplementary Figure 6). The short-term impact of PTG and resolution of symptoms and functional impairment can be seen at 12 to 24 post-operative months.

## Discussion

In this study, PTG performed within the context of a specialist service was associated with low peri-operative morbidity and excellent long-term oncologic outcomes. Major post-operative complications occurred in <2%, there were no postoperative mortalities and median length of stay was 8 days. The peri-operative outcomes compare favourably with previously reported series from high-volume centres, including those from National Institutes of Health and international multi-centre analyses, which reported major complication rates >10% and longer hospital stays.^17, 21^ The majority of patients had a pathological stage of pT1aN0M0 (88.0%), no lymph node metastases and complete gastric mucosal resection achieved for all. Our follow-up demonstrated DSS of 100% with median follow-up of 9.7 years. ^33^ These results demonstrated PTG undertaken within a specialist programme with expert endoscopic surveillance can be a safe curative treatment.

Weight loss was universal following PTG, with the most rapid decline occurring within the first 3 postoperative months and a nadir between 12 and 24 months. Total weight loss ranged from 15% to 20% of pre-operative body weight, with greater losses observed among pre-operative BMI >30 group. Partial weight re-gain occurred 2 to 5 years post-operatively among patients with normal pre-operative BMI. These findings are similar to those from Memorial Sloan Kettering Cancer Center cohort with the greatest loss seen in the pre-operatively obese group at a median of 28% at 2 years followed by 18% and 14% amongst overweight and normal weight groups respectively.^18^ We predict these observations may in part be attributed to the specialist dietetic input and regular follow-up of their nutritional status and weight with intervention for those with persistent nutritional compromise.

Health-related QOL declined across multiple symptom and functional domains in the early postoperative period, particularly within the first 3 months. Appetite loss, eating difficulties, fatigue, nausea, vomiting, and taste disturbance were most prominent. QOL measures returned to baseline within 6 to 12 months and remained stable for more than a decade, providing reassurance regarding long-term functional recovery. Patients managed with endoscopic surveillance alone demonstrated stable QOL over time. Among patients who underwent surveillance followed by PTG, serial assessments showed recovery of symptoms and function to near-baseline levels. Our finding that the recovery of symptoms and functional outcomes is preserved over a decade is novel and should provide some reassurance to patients with HDGC contemplating PTG. Notably, anxiety remained prominent among patients undergoing surveillance whereas improvement was observed following PTG. This finding may reflect reassurance associated with elimination of gastric cancer risk and highlights the need for psychological support for individuals undergoing surveillance.

We next assessed contribution of gender, age at surgery and preservation of the posterior vagal nerve on outcomes. Patients aged 35 or younger had better role functioning in line with the expectation that younger patients generally can recover over a shorter duration and return to daily activities. It is possible these patients were inclined to undergo PTG and it was based on patient choice. We demonstrated men experienced greater loss of appetite compared to women. Our results did not show a significant difference in QOL outcomes between vagal-preserving and non vagal-preserving surgery. The small numbers of patients with a complete vagotomy in our study may limit the power to detect a clinical difference and we did not routinely use intra-operative nerve monitoring to confirm posterior trunk was functional.^31^ Future work will be needed to explore the neurohormonal consequences of posterior vagotomy on patients.

Our study is limited by a relatively small sample size. Nonetheless, HDGC is a rare condition and larger cohorts are from MSKCC and NIH groups^17, 18^ A further limitation is voluntary reporting and it is possible we missed an important group with a negative experience who did not complete the questionnaires. Regular contact and close follow-up suggest this is less likely. We also facilitated questionnaire completion by online and paper formats. Lastly, our outcomes are in the context of a specialised service which may limit generalisability.

The authors propose this study improves our understanding of the long-term consequences of PTG. A strength of this study is that the effect of total gastrectomy on symptoms and function has been determined independent of a confounding disease process or concurrent systemic treatment for cancer. Identification of individual domains which had worse outcomes in specific sub-groups of patients will allow them to be targeted specifically to facilitate recovery and understand the physiological basis for this in future work. These findings are relevant to the wider literature as with improving oncological treatments and earlier detection of gastric cancer, the functional impact and long-term consequences become more important.

In conclusion, our study has shown PTG to be safe with excellent clinical and pathological outcomes. Despite the potential morbidity, we have shown that patients are able to achieve comparable outcomes to their baseline and to patients in surveillance within 12 to 24 months maintained in the long-term. These findings form a useful resource for patients contemplating PTG to understand medium and long-term consequences of PTG.

## Data Availability

All data produced in the present study are available upon reasonable request to the authors.

## Declarations

### Ethics approval and consent to participate

Consent was obtained prospectively from all and this study was carried out under the approval of Anglia and Oxford Medical Research Ethics Committee with reference number 97/5/32 and any amendments approved by the Cambridgeshire Research Ethics Committee

### Consent for publication

Not applicable.

### Availability of data and materials

Data are available from the corresponding author on reasonable request.

### Authors’ contributions

Hui Jun Lim, Claire Lamb, Sophie Samra, Nyarai Chinyama, Maria O’Donovan, Richard H Hardwick and J Robert O’Neill assisted with participant recruitment and data collection. Paul C Fletcher and Hisham Ziauddeen facilitated the set-up of data collection. Ju Yi Tai and Patrick Murphy aided in data collection. Lim HJ and O’Neill JR performed data analyses and wrote the manuscript. Di Pietro M and Fitzgerald R contributed intellectual guidance and examined the manuscript critically for publication. All authors have reviewed and approved the final manuscript.

## Acknowledgements

We would like to thank Marc Tischkowtiz and Samantha Grimes for their contributions and involvement in the care and follow-up of patients. In addition, we would like to thank Nikos Demiris for his guidance in the statistical analyses.

---

**Supplementary Figure 1:** Disease-specific Survival of Patients with Hereditary Diffuse Gastric Cancer Patients undergoing Prophylactic Total Gastrectomy.

**Supplementary Figure 2:** Effect of Age at Surgery on Change in Quality of Life Scores following Prophylactic Total Gastrectomy. Points represent mean ± 95% confidence interval. Age at surgery was grouped as ≤35 years old or >35 years old to allow two similarly sized groups. The outcome highlighted in the red box indicates a significant difference in percentage change from baseline scores between age-groups following surgery.

**Supplementary Figure 3:** Effect of Gender on Change in Quality of Life Scores following Prophylactic Total Gastrectomy. Points represent mean ± 95% confidence interval. The outcome highlighted in the red box indicates a significant difference in percentage change from baseline scores between genders.

**Supplementary Figure 4:** The Effect of Preservation of the Posterior Vagus on Change in Quality of Life Scores following Prophylactic Total Gastrectomy. Points represent mean ± 95% confidence interval.

**Supplementary Figure 5:** Summary of Impact of Prophylactic Total Gastrectomy on Health-related Quality of Life over Time.

**Supplementary Figure 6:** Comparison of Percentage Change in EORTC Scores (mean ± 95% confidence interval) for 3 Patients who underwent Endoscopic Surveillance followed by Prophylactic Total Gastrectomy.

