## Supplementary figures and images for "Long-term Outcomes of Prophylactic Total Gastrectomy in Patients with Hereditary Diffuse Gastric Cancer"

### Supplementary Figure 1

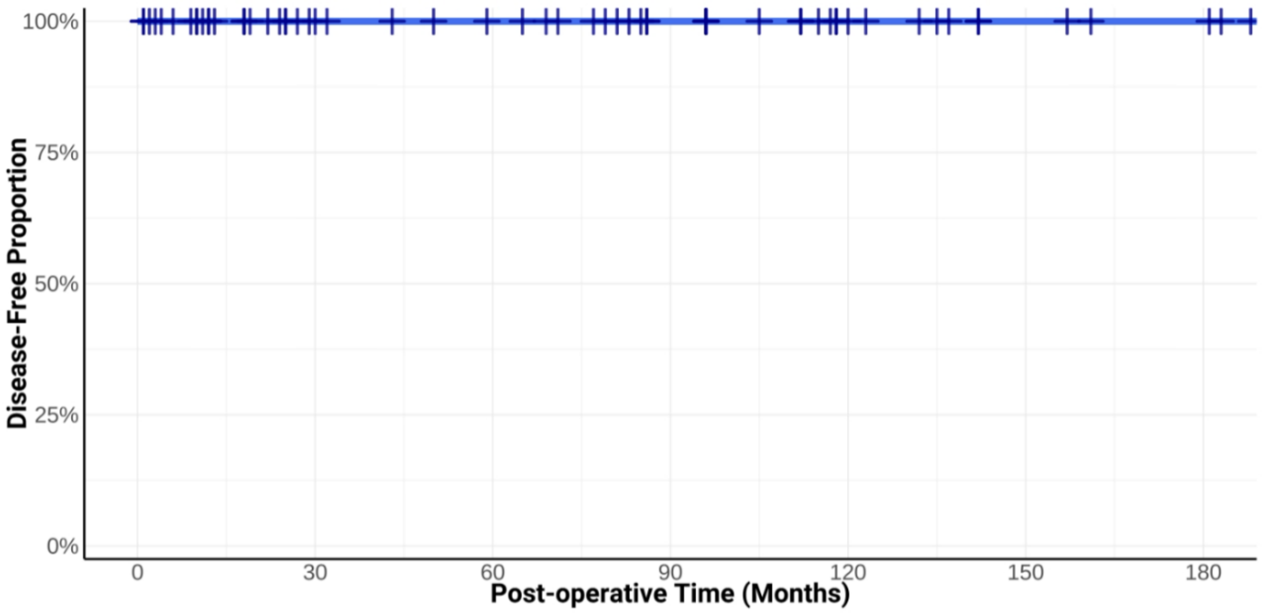


| | | | | | | | |
|---|---|---|---|---|---|---|---|
| **No. At Risk** | 60 | 38 | 33 | 23 | 13 | 6 | 4 |

### Supplementary Figure 2

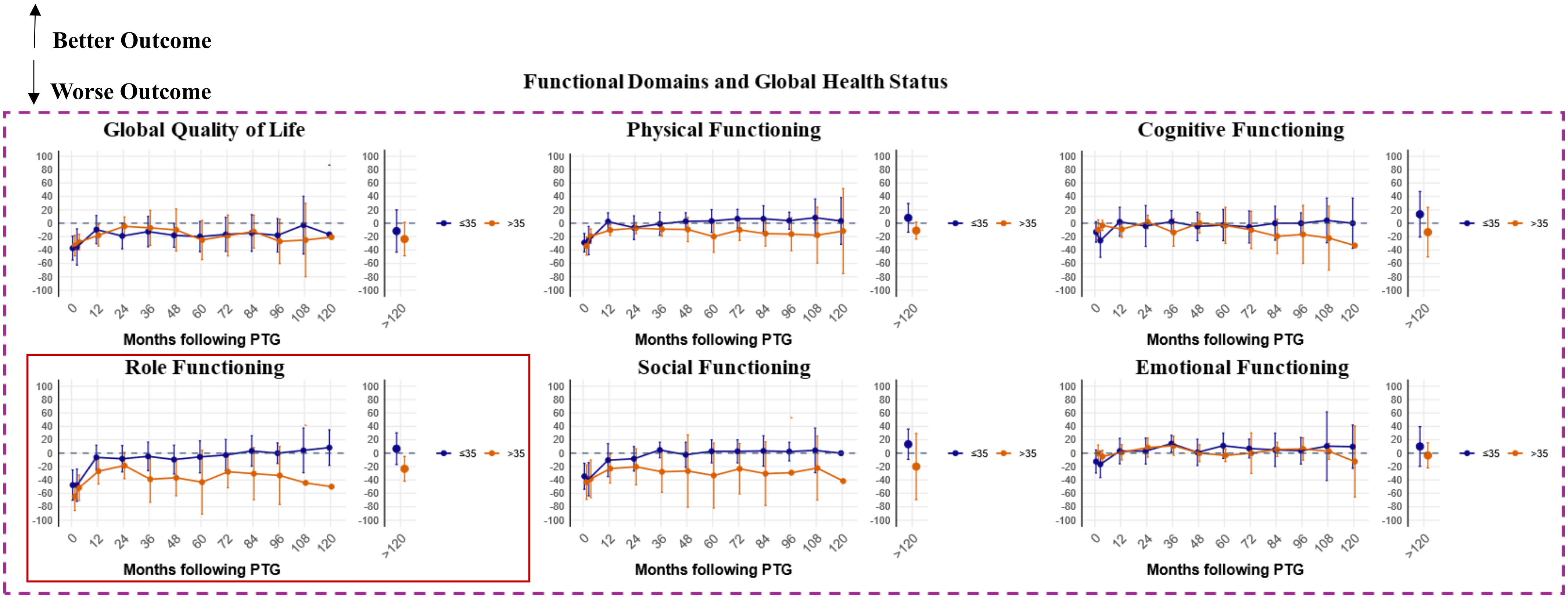

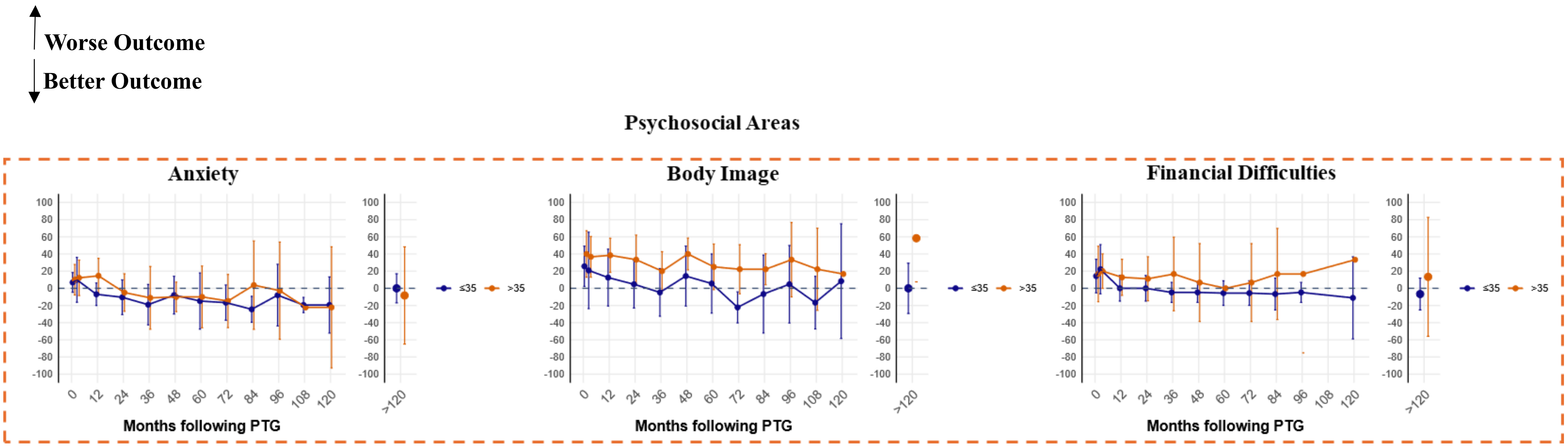

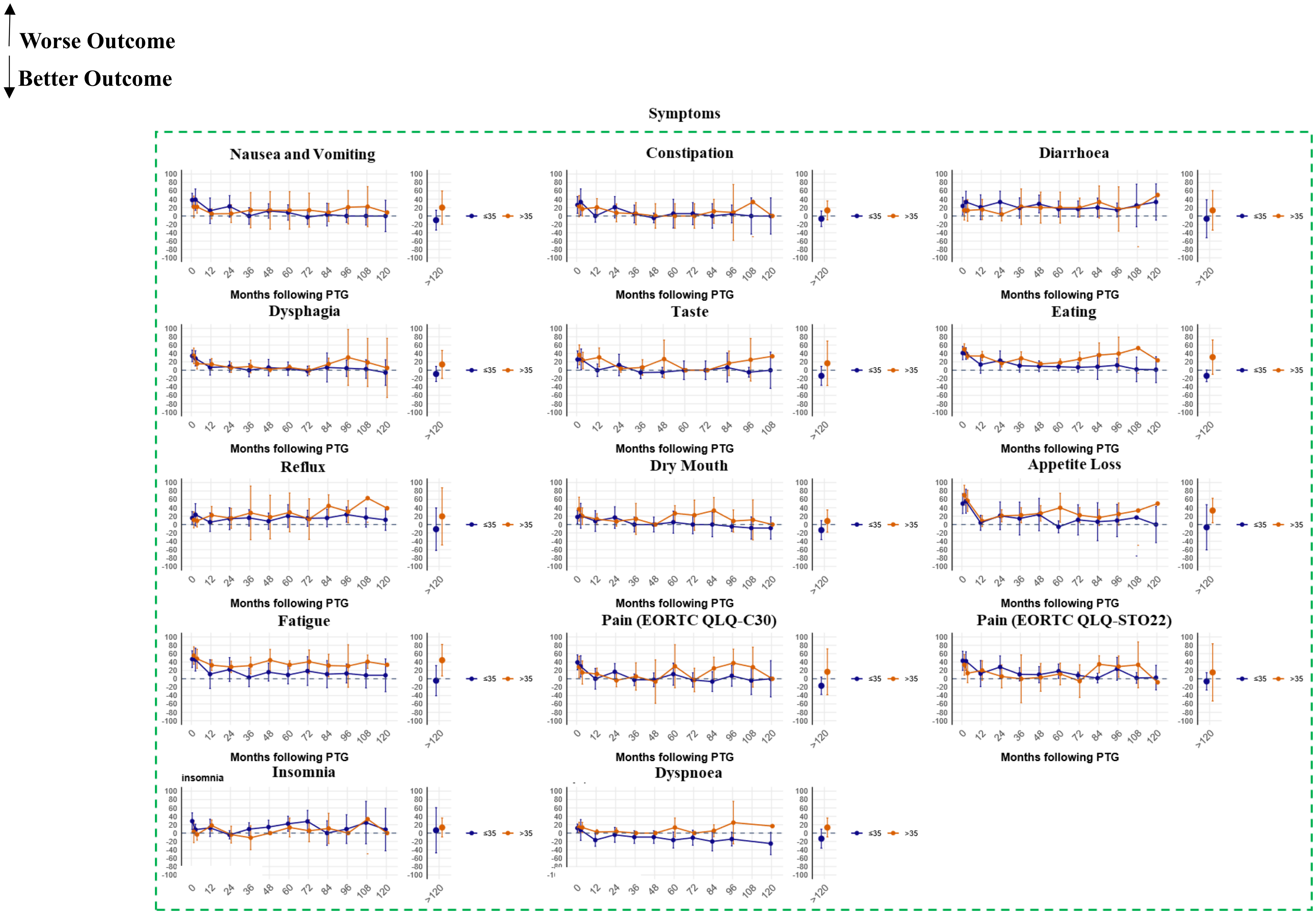

### Supplementary Figure 3

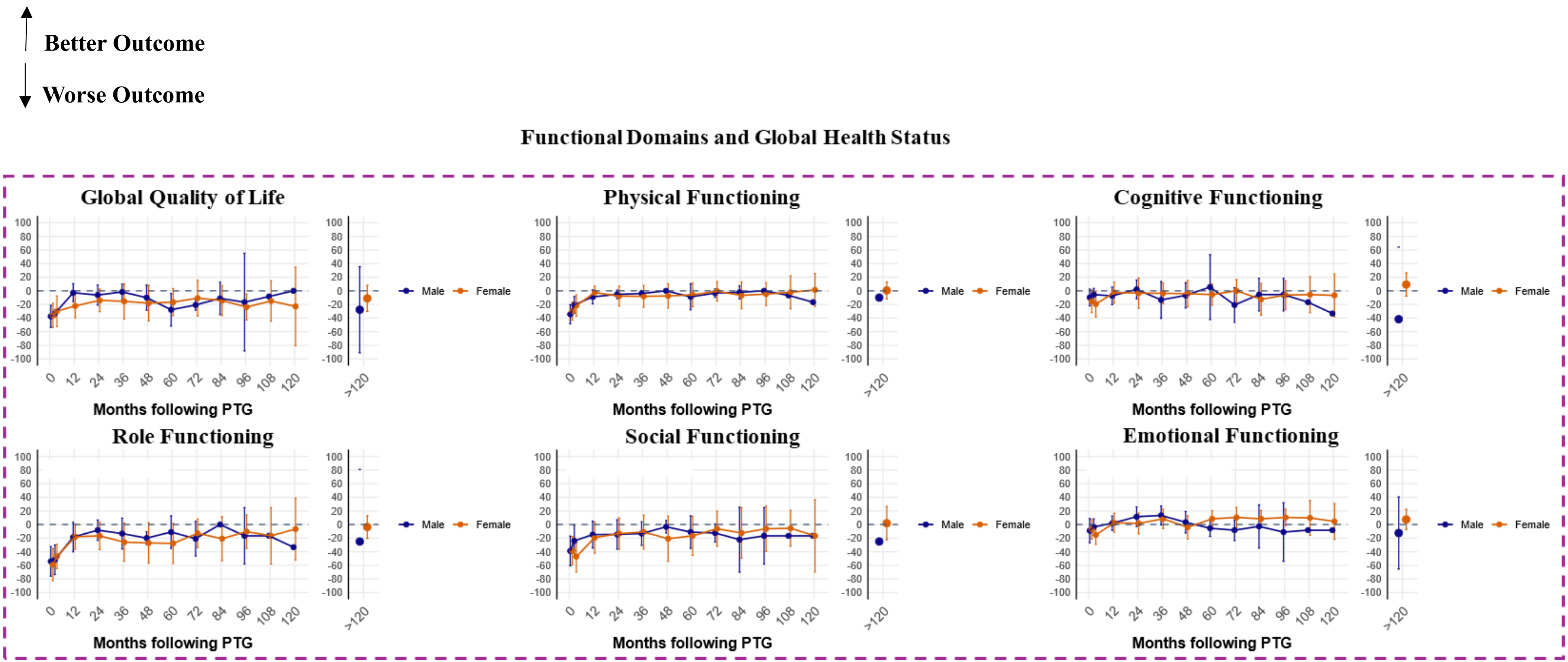

**Worse Outcome**

**Better Outcome**

#### Psychosocial Areas

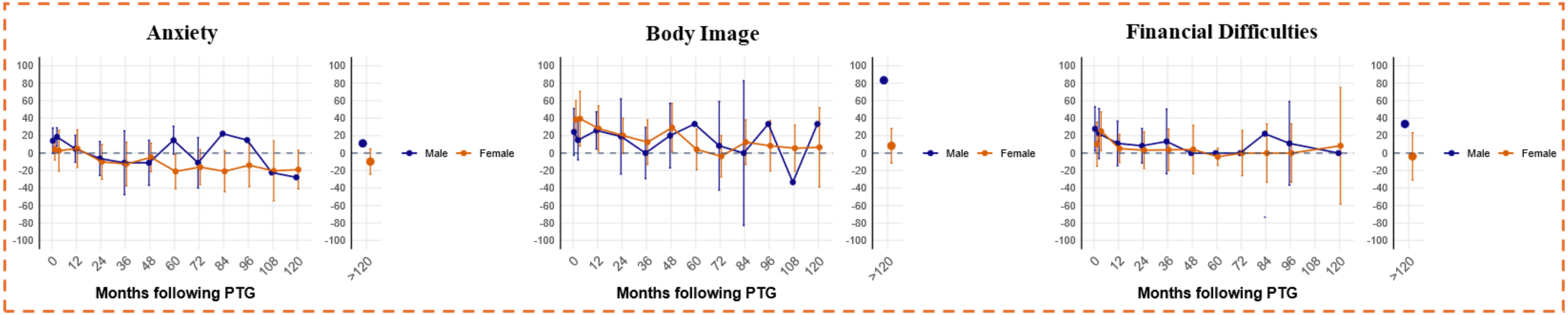

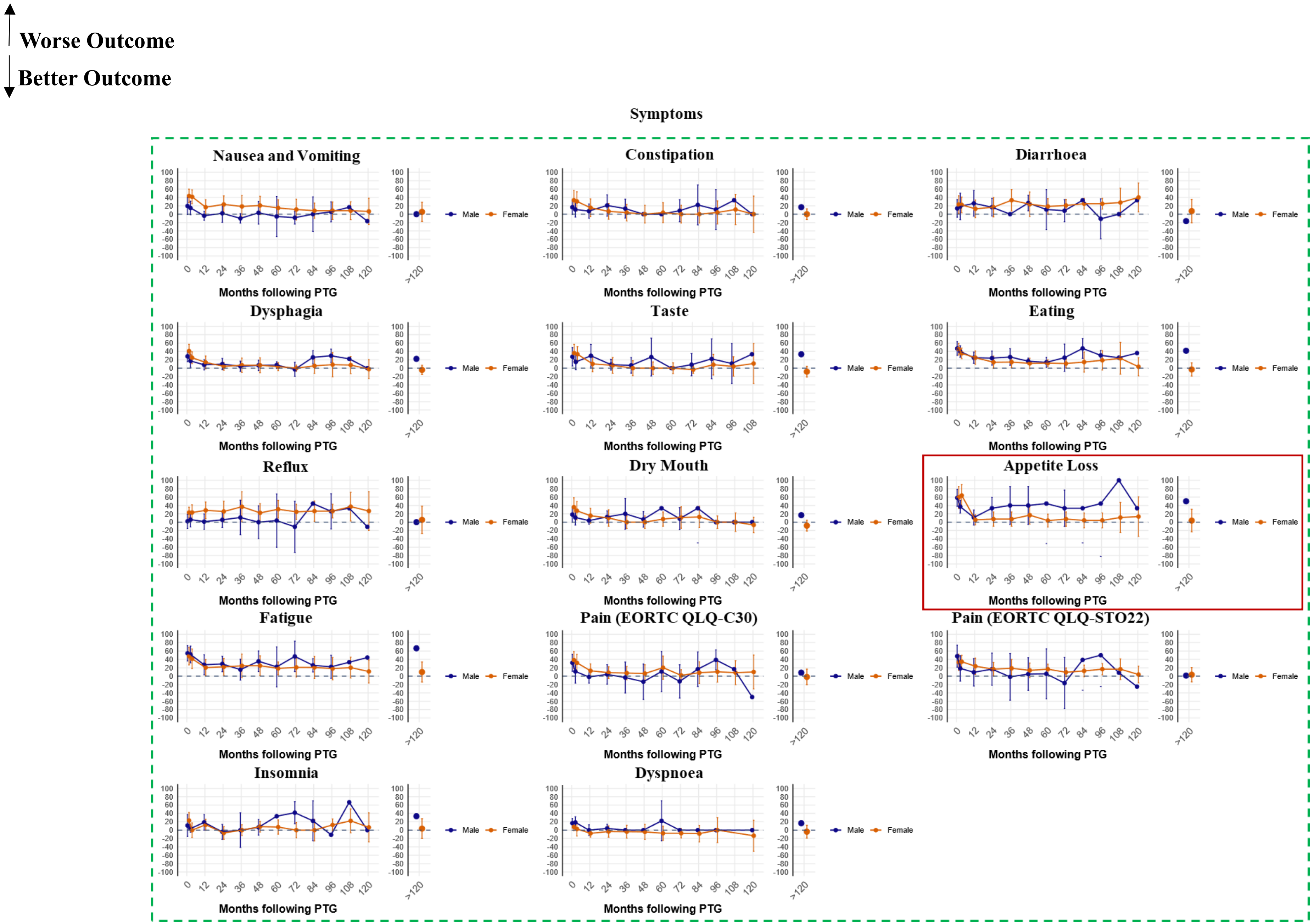

### Supplementary Figure 4

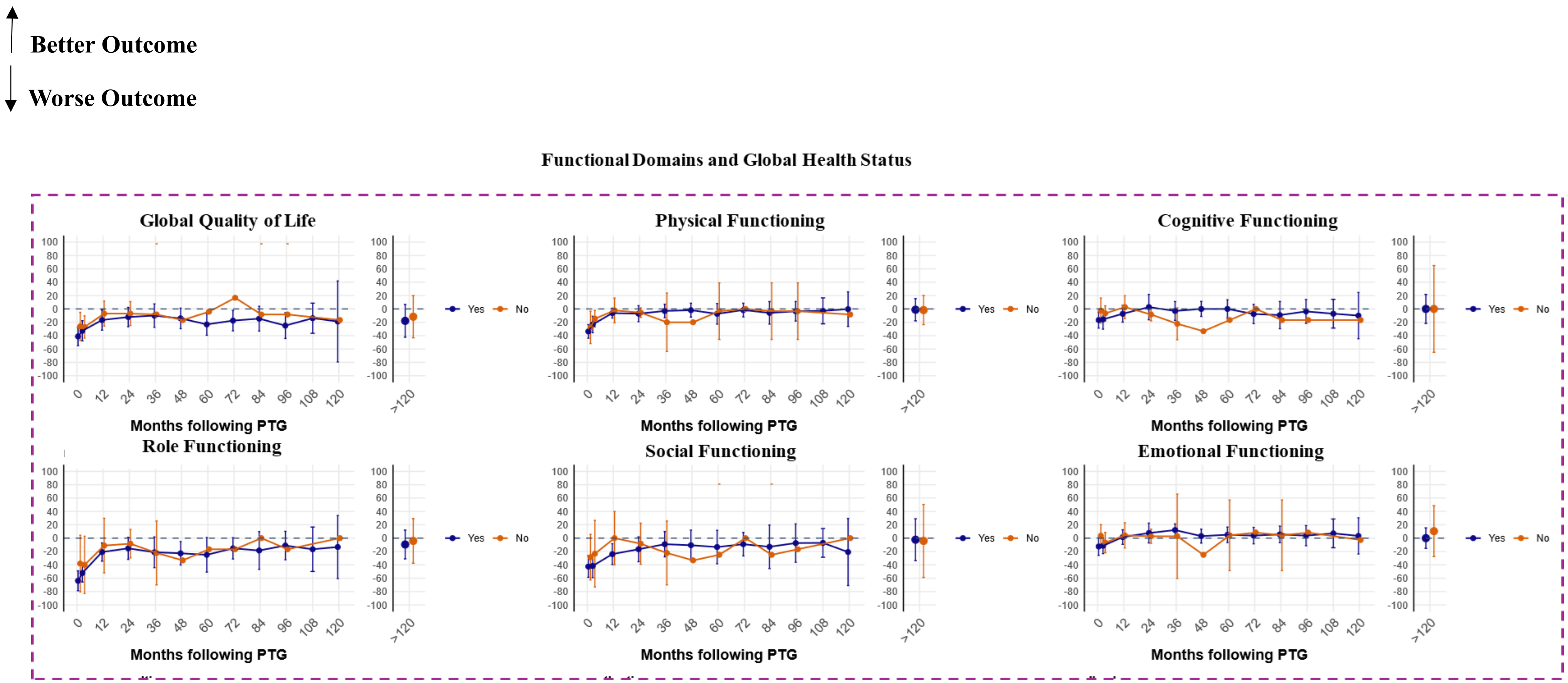

↑ **Worse Outcome**

↓ **Better Outcome**

**Psychosocial Areas**

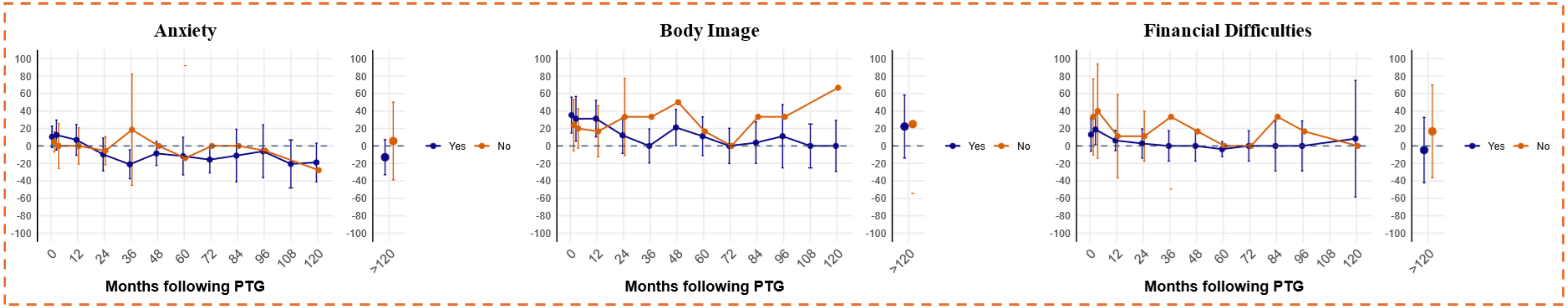

Worse Outcome

Better Outcome

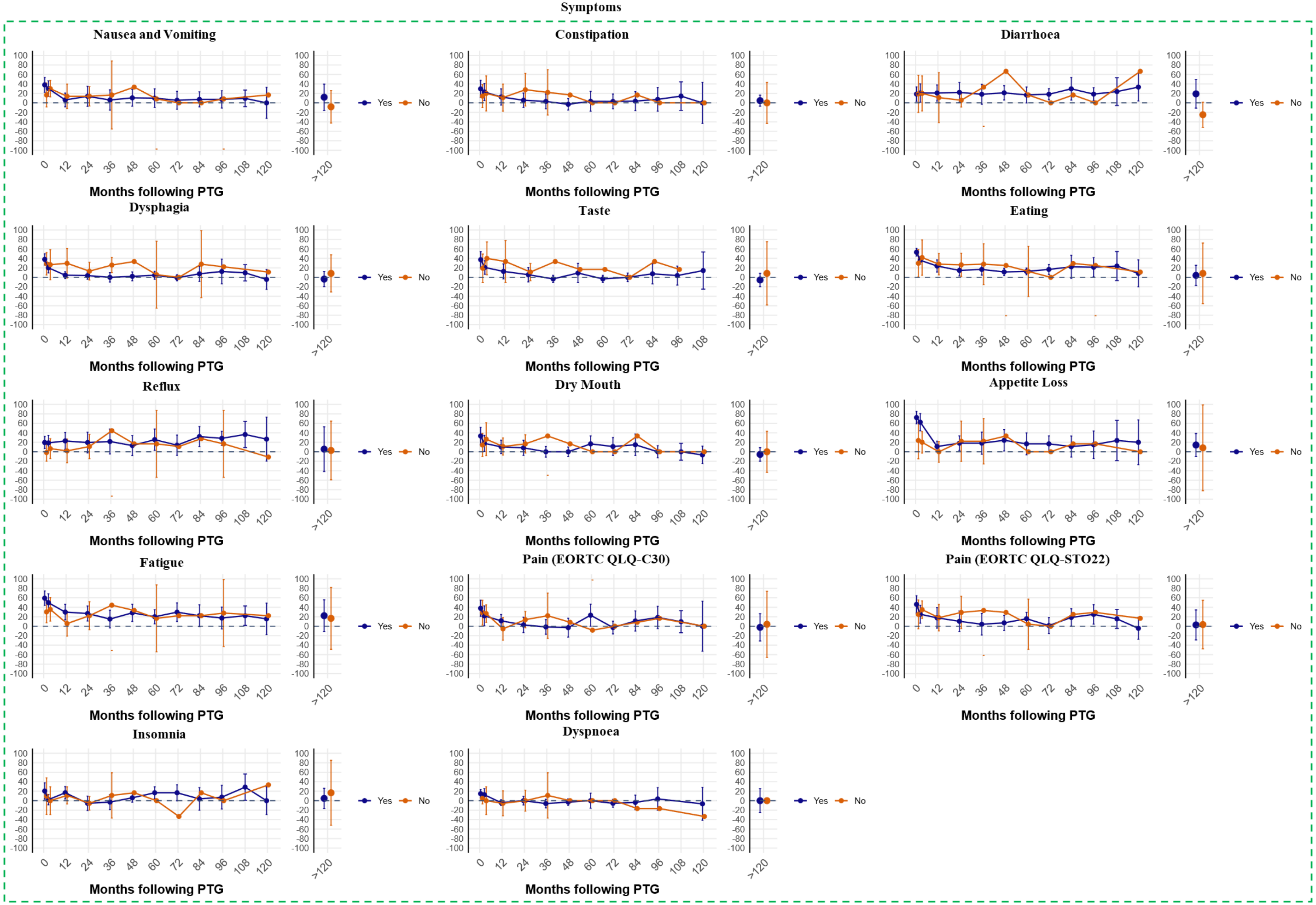

### Supplementary Figure 5

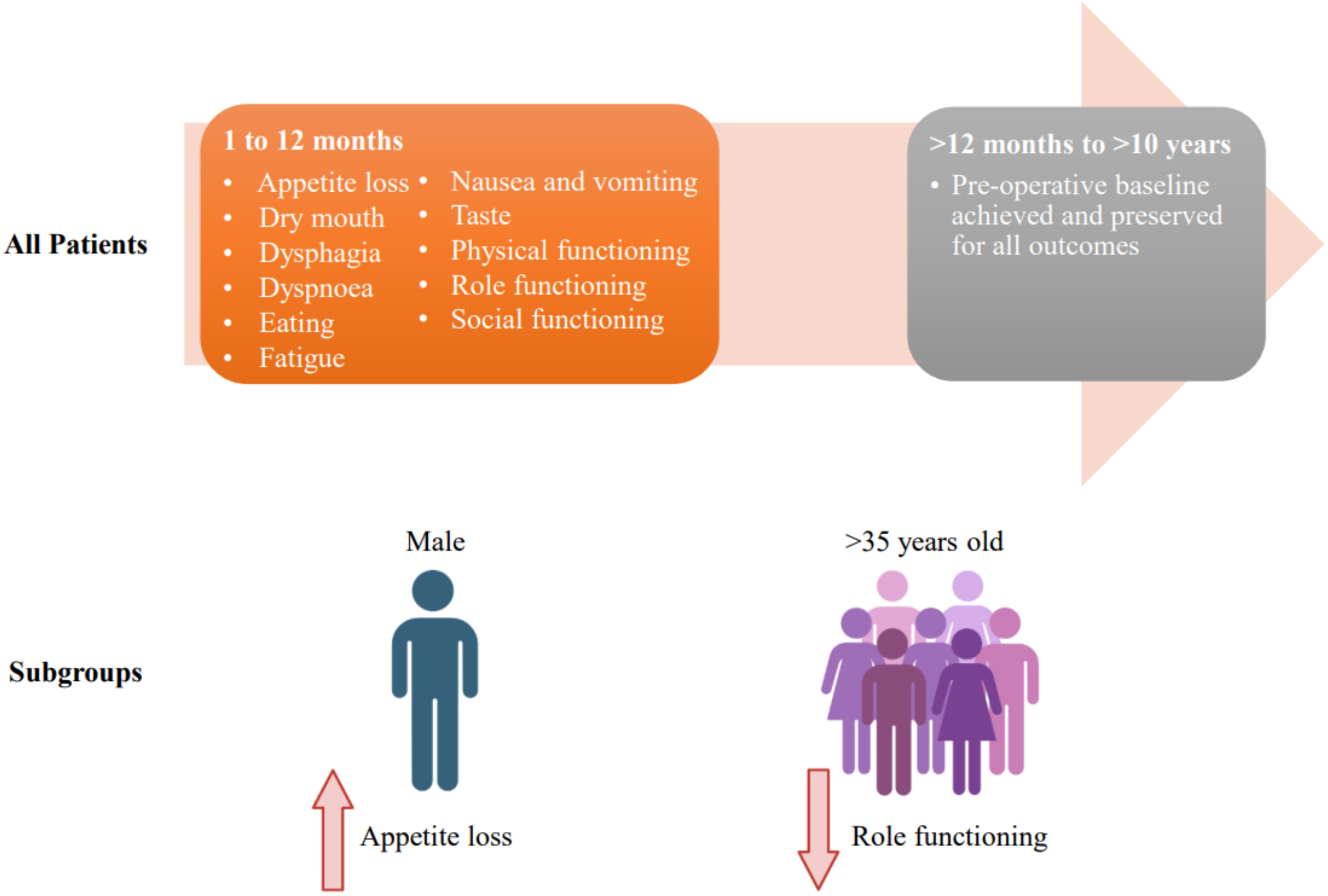

### Supplementary Figure 6

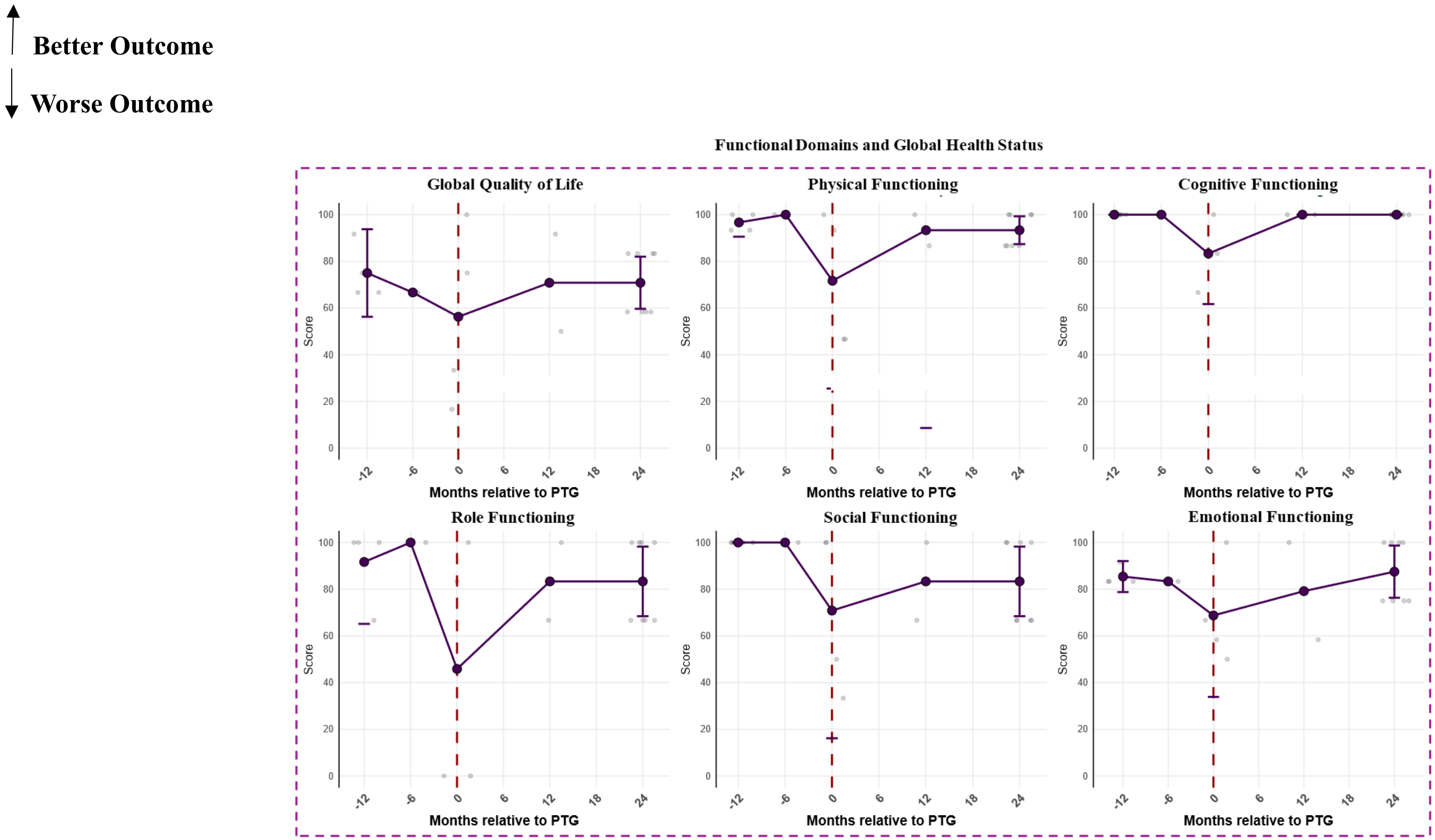

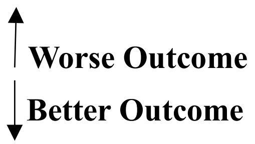


Psychosocial Areas

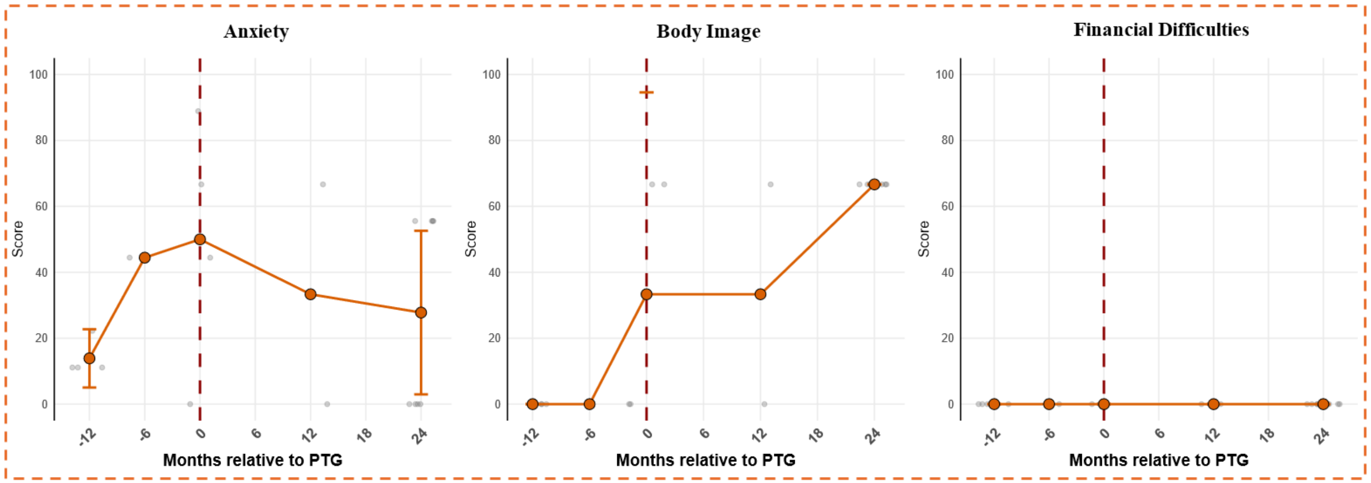

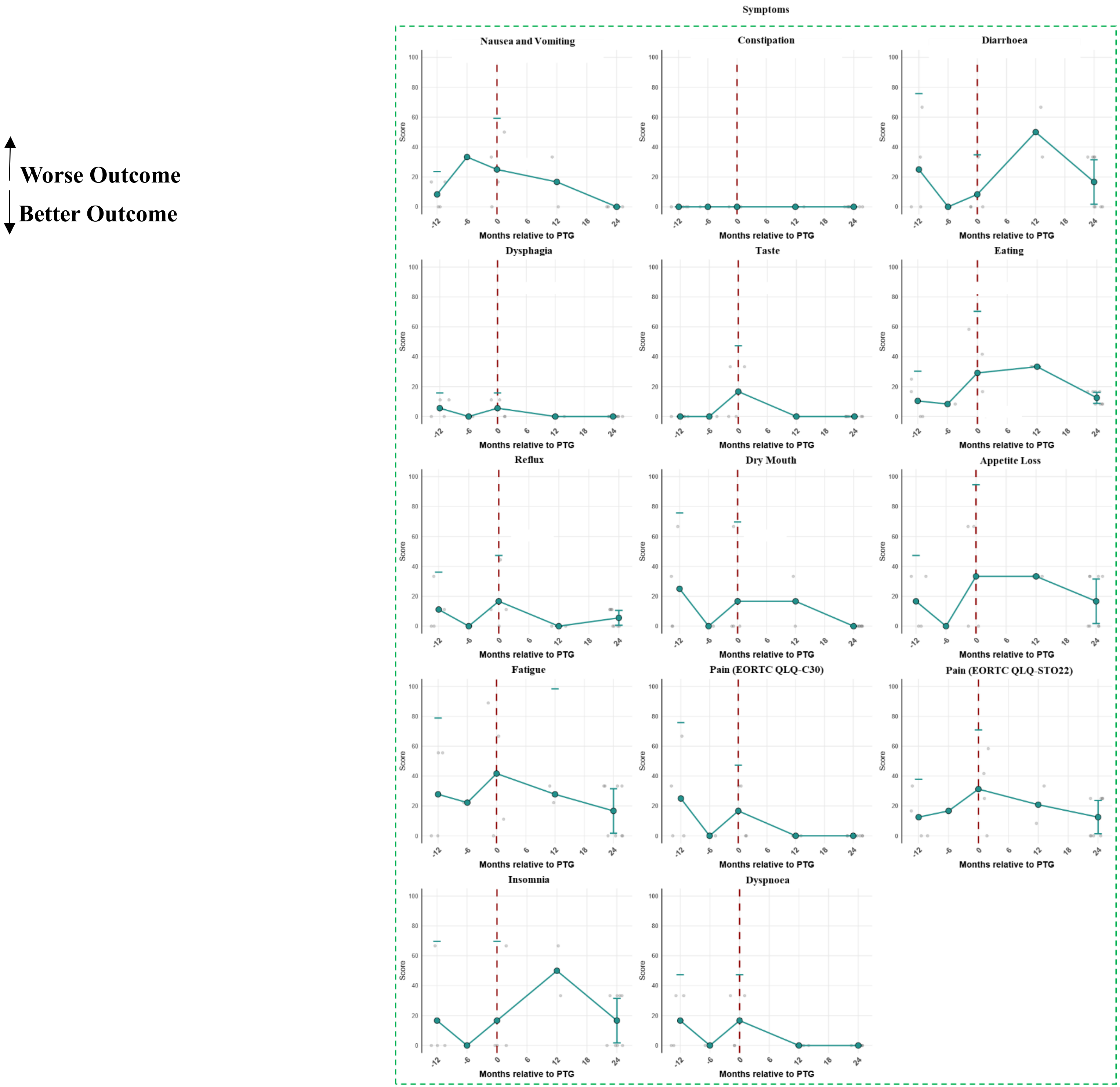
